# Detection of SARS-CoV-2 variant 501Y.V2 in Comoros Islands in January 2021

**DOI:** 10.1101/2021.04.08.21254321

**Authors:** Charles N. Agoti, George Githinji, Khadija S. Mohammed, Arnold L. Lambisia, Zaydah R. de Laurent, Maureen W. Mburu, Edidah M. Ong’era, John M. Morobe, Edward Otieno, Hamza Abdou Azali, Kamal Said Abdallah, Abdoulaye Diarra, Ali Ahmed Yahaya, Dratibi Fred Athanasius, Nicksy Gumede Moeletsi, Benjamin Tsofa, Philip Bejon, Peter Borus, D. James Nokes, Isabella Ochola-Oyier

**Author notes:** Correspondence /. Authors contributed equally.

## Abstract

Phylogenetic analysis of six SARS-CoV-2 genomes collected from the Comoros islands confirmed local circulation of the 501Y.V2 variant of concern during the country’s first major SARS-CoV-2 wave in January 2021. These findings demonstrate the importance of SARS-CoV-2 genomic surveillance and have implications for ongoing COVID-19 control strategies on the islands.

**Article summary line:** Circulation of SARS-CoV-2 501Y.V2 variant of concern in the Comoros Islands during a major COVID-19 infection wave in January 2021

## Main text

Although Comoros, an island country in the Indian ocean, detected its first case of SARS-CoV-2 on 30^th^ April 2020, it experienced its first major SARS-CoV-2 outbreak in January 2021 i.e., 10 months later (1). By 28^th^ February 2021, Comoros had 3,571 laboratory-confirmed SARS-CoV-2 infections, 2,748 (76.9%) of which were confirmed after 1^st^ January 2021.

Towards the end of 2020, in widely different geographical locations globally, three SARS-CoV-2 variants of concern (501Y.V1, 501Y.V2 and 501Y.V3) emerged that appeared to be considerably more transmissible and with potential to facilitate immune escape or cause more severe disease than the prior SARS-CoV-2 variants (2-4). The three variants possessed several defining amino acid changes, most of them occurring within the spike (S) protein (5). The S protein contains the domain that binds the virus to the human host cell receptor and is a key target for several vaccines (6).

## The study

To investigate if the variants of concern had a role in the rising number of SARS-CoV-2 cases in the Comoros in January 2021, 11 positive nasopharyngeal/oropharyngeal swab samples were sent to KEMRI-Wellcome Trust Programme (KWTRP) in Kilifi, Kenya, for genome analysis. KWTRP is one of the 12 designated WHO-AFRO /Africa-CDC specialized and regional reference laboratories for SARS-CoV-2 sequencing in Africa (7). The samples had been collected between 5^th^ and 11^th^ January 2021 from two islands (Ngazidja and Mohéli), **Table**. On receiving the samples on the 16^th^ and 17^th^ January 2021, viral RNA was extracted using the QIAamp Viral RNA Mini kit following the manufacturer’s instructions and analysed using the Sansure Biotech Novel Coronavirus (2019-nCoV) Nucleic acid Diagnostic real-time RT-PCR commercial kit which targets the nucleocapsid (N) and ORF1ab regions. Nine of the 11 samples were confirmed as SARS-CoV-2 positive by both gene targets (cycle threshold (Ct) <38.0). We proceeded to sequence six samples that had a Ct value of ≤ 29.0. The RNA was first reverse transcribed using the LunaScript® RT SuperMix Kit then amplified using the Q5® Hot Start High-Fidelity 2X Master Mix along with the ARTIC nCoV-2019 version 3 primers (8). The resultant amplicons were taken forward for library preparation and MinION (Mk1B) sequencing. The six samples were processed alongside 17 other samples from coastal Kenya to make a batch of 23 samples. The sequencing read-outs were demultiplexed and genomes assembled using a SARS-CoV-2 ARTIC Network Bioinformatics pipeline (8).

**Table.**
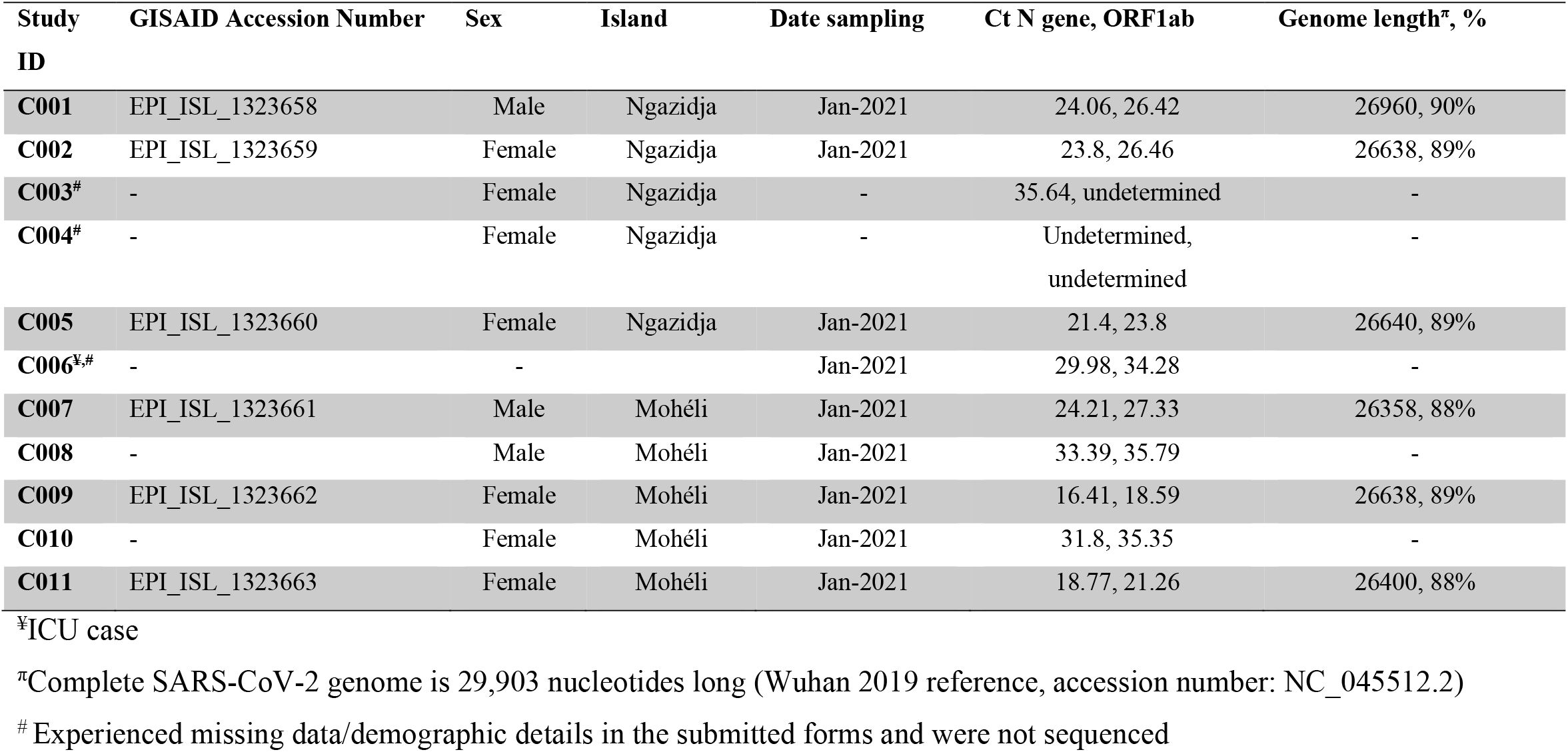
Baseline characteristics of the samples that were submitted to KEMRI-Wellcome Trust for sequencing and sequencing success.

We assembled >80% of the SARS-CoV-2 genome from each of the six sequenced samples, **Table**. The recovered genomes were classified into the lineage B.1.351 using the Pangolin toolkit v2.3.0 (9). The genomes possessed six of the eight variant 501Y.V2 defining amino acid changes in the S protein (i.e., L18F, D80A, D215G, K417N, D614G and A701V) plus a known three amino acid deletion at positions 243-245. Two additional defining amino acid changes (E484K and N501K) which were unconfirmed fell within a region that was not sequenced due to PCR amplicon drop-off. Our findings and confirmation of the presence of the SARS-CoV-2 501Y.V2 variant in Comoros samples was conveyed to Comoros authorities on the 22^nd^ January via the WHO-AFRO office to inform public health actions.

The six Comoros sequences differed only at three nucleotide positions: A13192G (1 genome), T23560C (2 genomes) and G27505T (1 genome). Compared to the Wuhan 2019 reference (Accession number: NC_45512.2), the Comoros genomes had 21-22 nucleotide substitutions that translated into 16-17 amino acid changes. We retrieved 75 random 501Y.V2 variant sequences deposited in GISAID and aligned them with the Comoros genomes using MAFFT v.7.313. A time-scaled phylogenetic tree of these sequences reconstructed in BEAST v 1.10.4 revealed that the Comoros genomes formed a monophyletic group together with genomes from Mayotte and France (**Figure**). This group diverged into 2 sub-clusters with the most recent common ancestor dated 30^th^ Oct-2020 (95%CI: 06^th^ Sep-2020 to 10^th^ Dec-2020).

**Figure.**
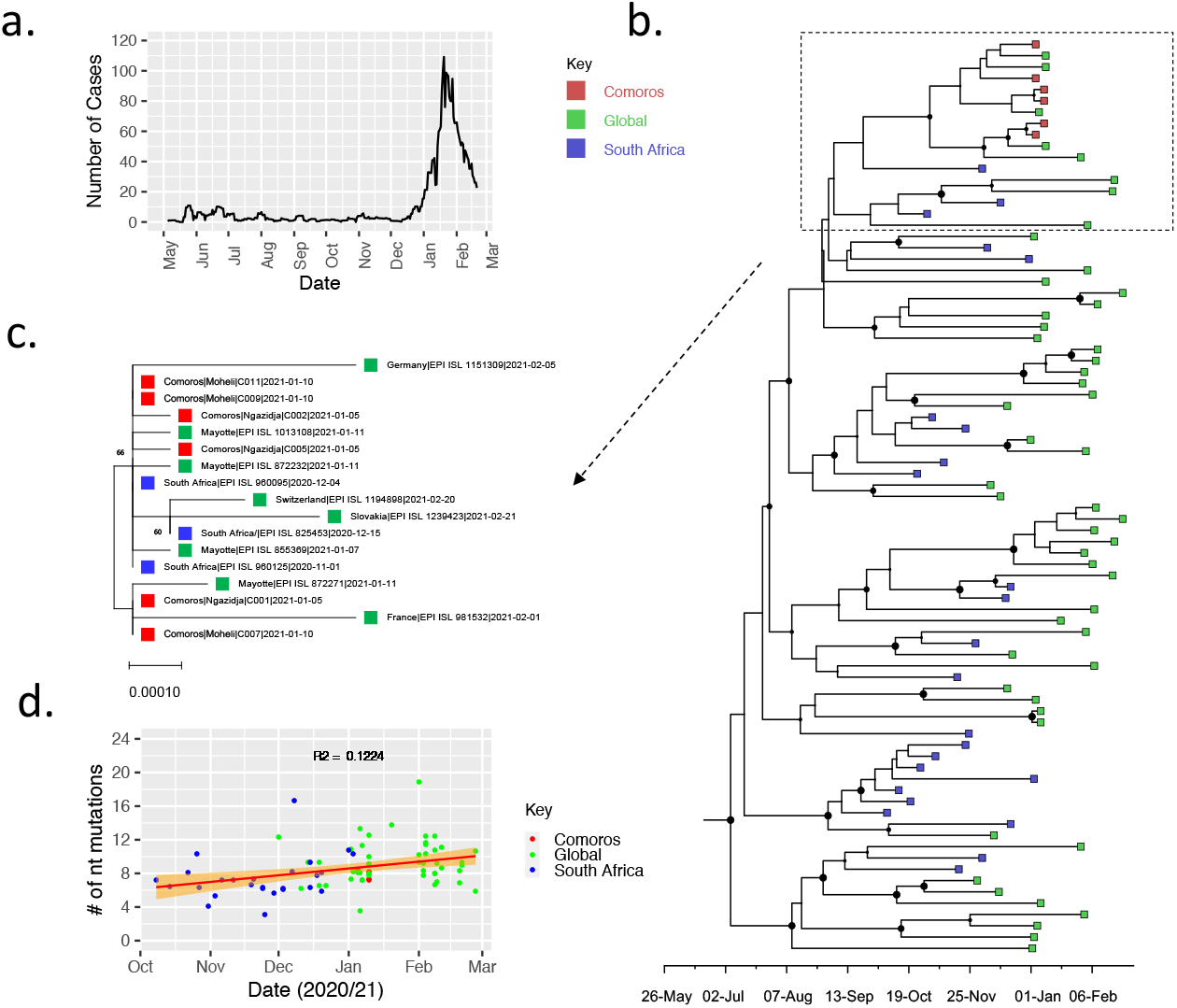
Genomic diversity of SARS-CoV-2 in Comoros Islands. **a**. Smoothed daily number of cases reported in Comoros Islands since the observation of the first case on 30^th^ April 2020 up to 19^th^ February 2020. **b**. time-scaled phylogenetic tree showing the placement of the six samples from Comoros (coloured in red) relative to other 501Y.V2 variant sequences from around the globe coloured in green. Those from South Africa are coloured blue. d. a zoomed in Maximum Likelihood tree of the clade containing the newly sequenced Comoros genomes. c. Root-to-tip divergence among the analysed 81 501Y.V2 genomes.

## Conclusion

We provide evidence of circulation of the 501Y.V2 variant during the first major SARS-CoV-2 epidemic peak in the Comoros. The 501Y.V2 variant was first identified in South Africa and has been reported in 41 countries as of 19^th^ February 2021. Initial data suggest that this variant exhibits up to 6-fold reduction in neutralization activity by post-vaccination sera or convalescent sera from individuals infected by prior variants (10). Thus, finding this variant in Comoros is concerning since it has potential to overcome pre-existing immunity derived from natural infection or vaccination.

Understanding the extent of spread of this variant in Comoros is limited by the low number of cases sequenced. As of 28^th^ February, 2021, the number of new cases in Comoros islands had considerably declined after peaking in mid-January (1). Comparison of the Comoros genomes with genomes from across the globe found a close relation with those from the neighbouring Mayotte. Our study demonstrates how continental genomic surveillance using the Sequencing Laboratory Network for Covid-19 can be utilised to inform response to the SARS-CoV-2 pandemic.

## Data Availability

The six genomes have been deposited in GISAID (Accession numbers EPI_ISL_1323658 - EPI_ISL_1323663).

## Ethical statement

The genomes generated were part of a regional collaborative COVID-19 public health rapid response. This manuscript therefore provides context on the genome data already available on GISAID. The whole genome sequencing study protocol was reviewed and approved by the Scientific and Ethics Review Committee (SERU) that sits at the Kenya Medical Research Institute (KEMRI) headquarters in Nairobi (SERU # 4035).

## Acknowledgements

We thank the WHO-AFRO and the WHO Kenya and Comoros country offices for facilitating sharing of the positive samples that were sequenced; Further, we thank all laboratories that have shared SARS-CoV-2 sequence data on GISAID (Appendix**). For the purpose of Open Access, the author has applied a CC-BY public copyright licence to any author accepted manuscript version arising from this submission**.

## First author biography

Dr Charles N. Agoti is a post-doctoral Molecular Virologist while Dr George Githinji is post-doctoral Bioinformatician, both are based at the KEMRI-Wellcome Trust Research Programme, Kilifi, Kenya.

